# Fixation beats – a novel marker for reaching the left bundle branch area during deep septal lead implantation

**DOI:** 10.1101/2020.10.04.20206573

**Authors:** Marek Jastrzębski, Grzegorz Kiełbasa, Paweł Moskal, Agnieszka Bednarek, Aleksander Kusiak, Tomasz Sondej, Adam Bednarski, Marek Rajzer, Pugazhendhi Vijayaraman

## Abstract

**Introduction:** One of the challenges of left bundle branch (LBB) pacing is to place the pacing lead deep enough in the septum to obtain capture of the LBB, yet not too deep to avoid perforation. We hypothesized that the occurrence of the ectopic beats of qR/rsR’ morphology in V1 lead (fixation beats) during the lead fixation would predict that the final desired intraseptal lead depth was just reached, while the lack of fixation beats would indicate too shallow position, and need for more lead rotations.

**Methods:** Consecutive patients during LBB pacing device implantation were analyzed retrospectively and then prospectively with respect to the occurrence of the fixation beats during each episode of lead rotation. We compared the presence of fixation beats during the lead rotation event directly before the LBB capture area depth was reached versus during the events before intermediate/unsuccessful positions.

**Results:** A total of 339 patients and 1278 lead rotation events were analyzed. In the retrospective phase, the fixation beats were observed in 327/339 of final lead positions and in 9/939 of intermediate lead positions (p<0.001). The sensitivity, specificity, positive and negative predictive value of the LBB area fixation beats as a marker for reaching the LBB capture area was 96.4%, 99.0%, 97.3% and 99.0%, respectively. In the prospective, fixation-beats-guided, implantation phase the fixation beats were observed in all patients and only at the LBB capture depth.

**Conclusions:** Monitoring fixation beats during deep septal lead deployment can facilitate the procedure and possibly increase the safety of lead implantation.

## Introduction

Left bundle branch (LBB) pacing is emerging as the most feasible method of conduction system pacing both for bradyarrythmia and heart failure treatment.^1-4^ Several aspects of this new pacing modality need to be better characterized. One of the challenges of this technique is to advance the LBB pacing lead intraseptaly deep enough to ensure capture of the left bundle branch, yet not too deep to avoid immediate or delayed perforation of the interventricular septum. During advancement of the LBB pacing lead ectopic ventricular beats are observed resulting from the irritation of tissues as the leads crosses the septum (Figure 1); some of these have relatively narrow QRS complexes of qR/rsR’ morphology in lead V1, very suggestive that they originate within LBB or its arborization. It was recently illustrated that these premature complexes can be exploited to guide LBB pacing procedure.^5^ We hypothesized that the occurrence of the LBB area ‘fixation beats’, as we dubbed them, would predict that the final desired intraseptal lead depth was reached, while the lack of LBB area fixation beats would indicate too shallow lead position, and need for more lead rotations.

**Figure 1.**
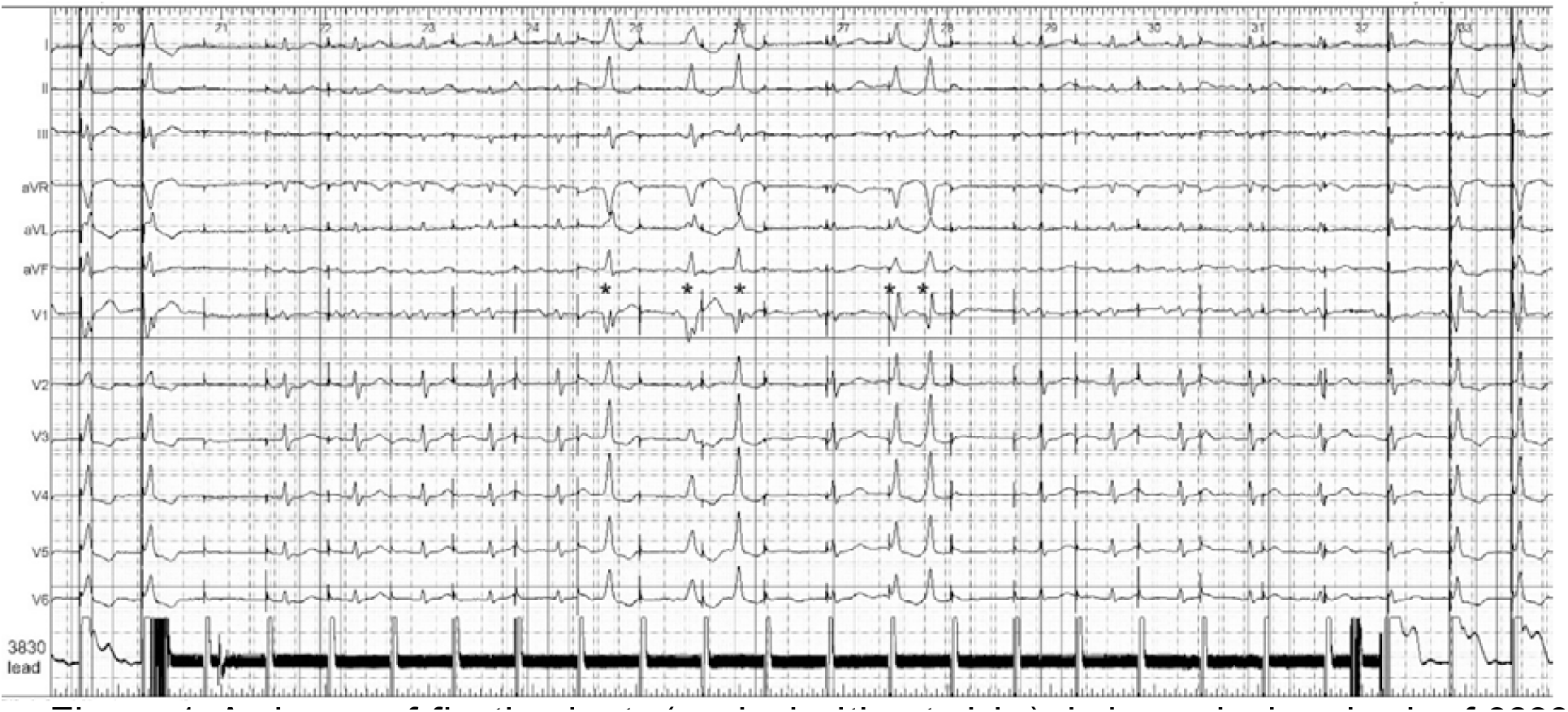
A shower of fixation beats (marked with asterisks) during a single episode of 3830 lead rotation for deep septal lead placement. During rotation, the lead was disengaged from the alligator clip connection to the external pacemaker (marked with the appearance of the noise signal on the 3830 lead channel) for 12 seconds. The first three fixation beats were classified as ‘deep septal fixation beats’ while the last two, of qR morphology, as ‘LBB area fixation beats’. The fixation beats showed progressive change of QRS morphology that is compatible with progressively deeper origin within the interventricular septum. Please note, that the paced QRS morphology when the lead was reconnected with the alligator clip/external pacemaker is nearly identical as the last LBB area fixation beat, while the first fixation beat morphology is nearly identical as the initial paced QRS that served to identify the optimal site for lead advancement. These observations indicate that the fixation beat morphology reflect the actual position of the screw-in electrode helix. LBB – left bundle branch.

The aim of the study was to investigate predictive value of LBB area fixation beats for reaching the final desired pacing lead position within the interventricular septum.

## Methods

### Study population

Consecutive patients who received permanent left bundle branch pacing devices were analyzed with respect to the occurrence of LBB area fixation beats during the lead advancement deep into the interventricular septum. In our electrophysiology laboratory every patient with indications for pacemaker or CRT implantation receives conduction system pacing based device. At the implanting physician’s discretion, LBB pacing device is chosen as the primary choice or as a secondary choice (after an attempt to obtain His bundle pacing). The fixation beat analysis was initially conducted in a retrospective fashion, and, once this method was adopted by us as a major tool guiding the LBB lead implantation, the analysis was performed in a prospective fashion. All other clinical and procedure related data were collected on the basis of our prospectively maintained conduction system pacing database.

### Fixation beats definition and analysis

Fixation beats were defined as ectopic ventricular beats occurring during the lead rotations for deep intraseptal deployment. These were categorized as **deep septal fixation beats** (similar or more narrow than the initially obtained septal paced QRS complexes, albeit, without qR / rsR’ morphology in lead V1 and > 130 ms in duration) and **LBB area fixation beats** (Qr/qR/rsR’ morphology present in lead V1 and/or QRS ≤ 130 ms) – see Figures 1 and 2.

**Figure 2.**
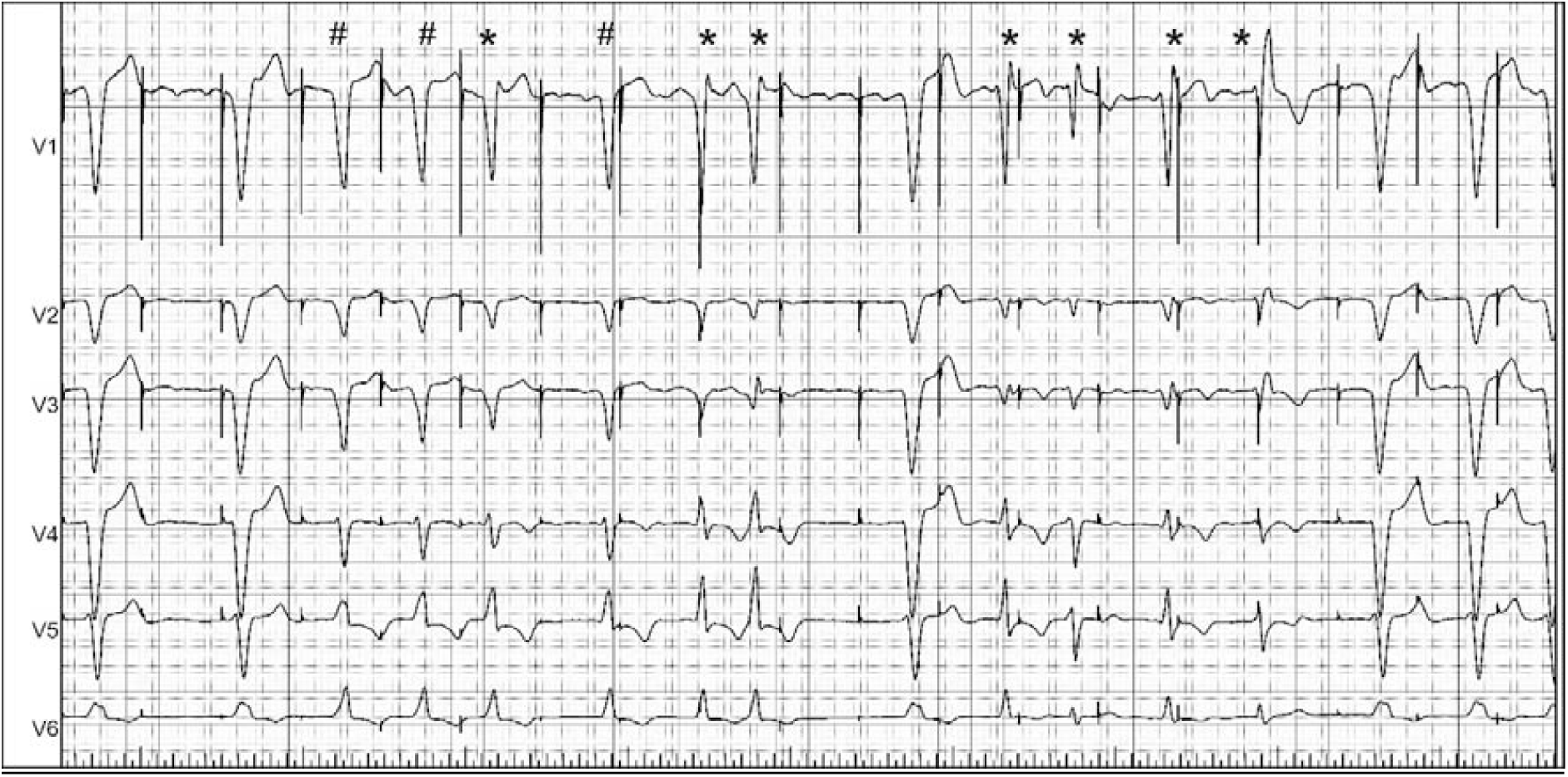
A shower of fixation beats (marked with asterisks and hashes) in a patient with preexistent LBBB during an episode of deep septal lead deployment. Note progressively more selective irritation of the LBB indicated by narrowing of fixation beat QRS and increase in the size of R wave in lead V1. The QRS complexes marked with hashes are likely fusion beats between supraventricular QRS and a fixation beat. LBBB – left bundle branch block. LBB – left bundle branch.

Both the 12-lead ECG and the endocardial electrogram from the SelectSecure™ 3830 pacing lead (Medtronic Inc, Minneapolis, MN, USA) were continuously recorded on an electrophysiology system (Lab System Pro, Boston Scientific, USA) during the whole implantation procedure. Each episode of the 3830 lead rotation during the procedure was easily identified, as it was marked by the appearance of a noise signal due to the disconnected alligator clip, interspersed with periods of pace-mapping for the assessment of paced QRS morphology, resulting in a very distinctive pattern in the electrophysiology system record (Supplemental Figure 1). All lead rotation episodes were analyzed with regard to the occurrence of fixation beats.

### Implantation procedure

For all procedures, only the 3830 model lead and delivery sheaths from the SelectSite™ family were used (Medtronic Inc., USA). The LBB-pacing device implantation generally followed the methods recently described by Huang, Vijayaraman and others, ^2, 3, 6^ albeit, with some potentially important modifications delineated below. The implantation site was chosen on the basis of the right ventricular septal pacemapping and the tricuspid summit position, no extensive search for His bundle potential was performed. The optimal paced QRS characteristics was considered to be present when there were R in lead II, and RS/rS/QS in lead III accompanied by QS with notched nadir in V1; other QRS morphologies suggestive of more superior (R in II and III) or more inferior (rS/QS in II and III) lead position or and/or lack of notch in V1 were accepted when the optimal pace-mapped QRS was not obtained or the sites with such QRS did not offer deep septal lead deployment/LBB capture. The summit of the tricuspid ring was approximated on the basis of the delivery sheath movement, atrial vs. ventricular endocardial signals, His bundle potential (if encountered) and occasionally contrast injection via the delivery sheath. The LBB target area was defined as approximately 2 cm apical from the tricuspid summit, extending conically 2 cm to the superior and inferior mid-septum (Figure 3). The initial deployment attempt consisted of approximately 5 rapid rotations, and then, in an interrupted fashion, pacemapping was performed from the 3830 lead. More lead rotations were added until a satisfactory paced QRS morphology was obtained and/or LBB capture was confirmed otherwise. When after the initial 5 rotations the torque created in the lead was not transmitted but remained in the lead body, the ‘entanglement response’ or fibrous site was suspected and the implantation site was changed.^7^ When after the initial 5 rotations or after the subsequently added rotations there was no progressive change in paced QRS morphology, the ‘drill effect’ was suspected and either more forward pressure was attempted, sheath angle/torque was modified to offer better support for the lead or the implantation site was changed.^7^

**Figure 3.**
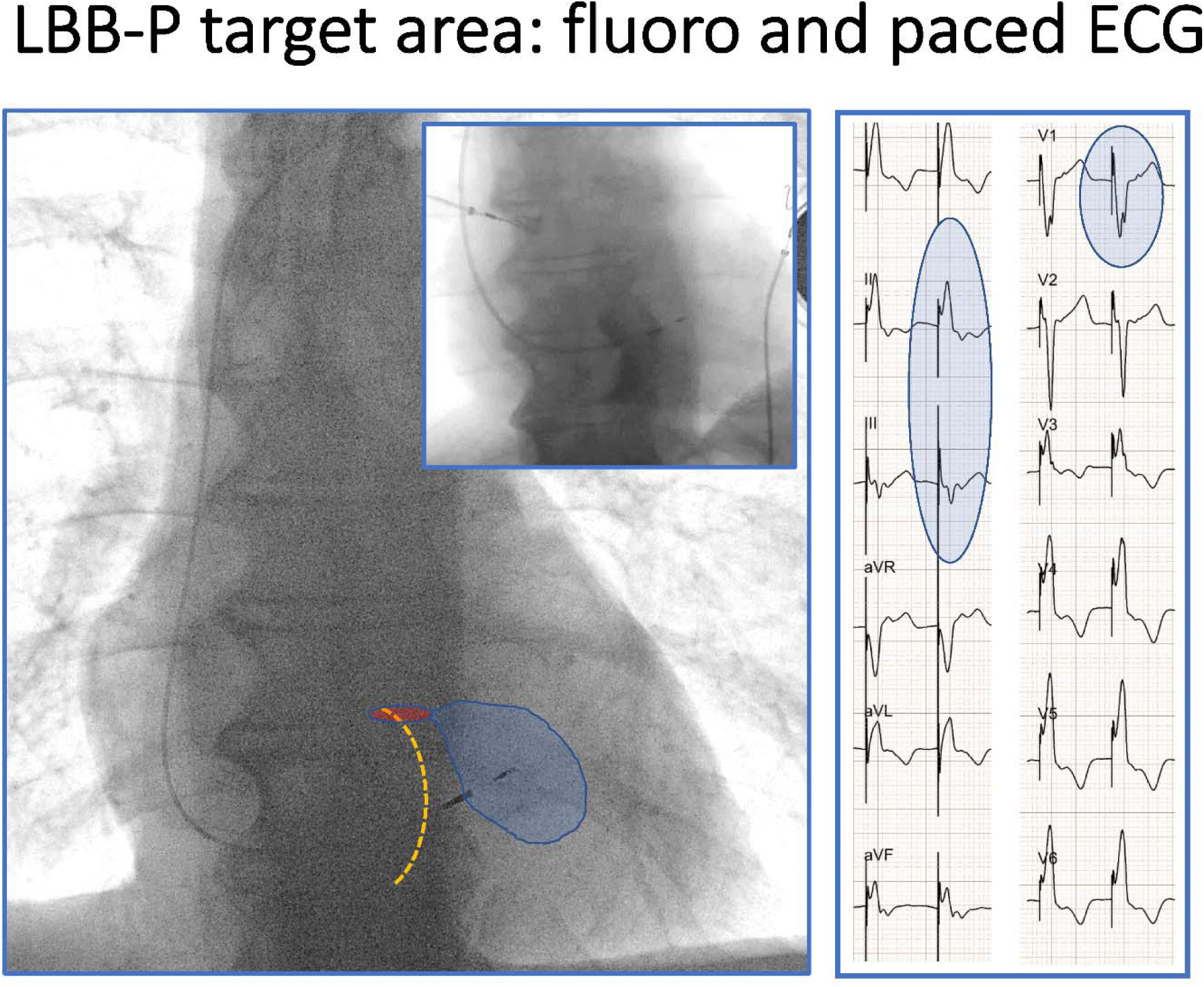
Schematic representation of the target area for deep septal lead deployment as used in the current study. Left bundle branch pacing (LBB-P) area, shaded in blue, was determined on the basis of the tricuspid ring position determined fluoroscopically and electrophysiologically and on the basis of the paced QRS morphology (positive QRS in II, negative QRS component in III and “W” shape in V1). Presumed His bundle area was shaded in red. Inset shows contrast enhanced determination of the tricuspid ring position

In the prospective, fixation-beat-guided, phase of the study the implantation technique was modified. Instead of using interrupted pace-mapping after each set of lead rotations, the initial lead rotations were continued (up to 10 rotations), with the lead disconnected until the LBB fixation beats appeared and then the lead rotations were immediately stopped and pace-mapping was performed to confirm whether the satisfactory, final lead position was reached (Figure 1; Supplemental Movie 1).

The deep septal paced QRS obtained during pace-mapping was considered satisfactory when the initial paced QRS morphology in V1 of QS type was transformed into Qr/QR or rsR’/rSr’ morphology (i.e. an R/r wave at the end of QRS appeared) or QRS narrowed to ≤ 120 ms (measured from the clear QRS onset in surface ECG, i.e. without the initial latency period). LBB capture was diagnosed when either direct proof was obtained with differential pacing output or programmed / burst stimulation or one of the indirect, arbitrary criteria was present (time to R wave peak in V5/V6 < 85 ms, QRS ≤ 120 ms, or LBB potential on the 3830 lead electrogram).^2, 6-8^

### Statistical analysis

Continuous variables are presented as means and standard deviations or medians and interquartile range (IQR) - for data with non-normal distribution, while categorical variables are presented as numbers and percentages. The occurrence of the fixation beats at successful sites inside the septum was compared with the occurrence of the fixation beats at unsuccessful depths/sites using chi-square. Predictive value of the fixation beats for reaching the final intraseptal lead position was assessed using standard measures: sensitivity, specificity, positive and negative predictive value. Groups were compared with Student t-test, Mann-Whitney U test or Fisher Exact Test. P value < 0.05 was considered significant. Statistical analysis was performed using StatSoft Statistica v.13.3 (TIBICO Software Inc, Palo Alto, California, USA).

## Results

### Study population, procedural characteristics, and fixation beats as diagnostic marker

Starting from the 12 June 2018 (first LBB pacing case in our institution) a total of 784 consecutive patients, that underwent pacemaker or CRT device implantation, were screened. Out of these, 358 patients received LBB pacing devices, the remaining received either His bundle devices (n=313), or were conduction system pacing failures, in case of which either classic biventricular CRT (n=36) or right ventricular myocardial pacing (n=59) device was implanted. After exclusion of 5.3% of LBB pacing cases (artefacts, file loss, alternative implantation technique, etc.) finally 339 LBB-paced patients were included into the further analysis (including the final 21 prospective cases). Basic clinical and implantation procedure related characteristics of the included patients are presented in Table 1.

**Table 1.** Basic characteristics of the studied group (n = 339)

|  |  |
| --- | --- |
| Age [years] | 76.1 ± 10.8 |
| Male gender | 180 (53.1%) |
| Pacing indication [n]: |  |
| • Sick sinus syndrome | 78 (23.0%) |
| • Atrioventricular block | 139 (41.0%) |
| • Atrial fibrillation with bradycardia | 38 (11.2%) |
| • Heart failure | 84 (24.8%) |
| Comorbidities [n]: |  |
| • Diabetes mellitus | 138 (40.7%) |
| • Coronary heart disease | 132 (38.9%) |
| • Heart failure | 153 (45.1%) |
| • Hypertension | 278 (82.0%) |
| • Severe valvular disease | 42 (12.4%) |
| Left ventricular ejection fraction [%] | 48.8 ± 14.9 |
| Left ventricular end-diastolic dimension [mm] | 50.9 ± 8.8 |
| Native QRS duration [ms] | 127.5 ± 34.1 |
| Native QRS type: |  |
| • RBBB | 25 (7.4%) |
| • RBBB + LAH/LPH | 52 (15.3%) |
| • LBBB | 42 (12.4%) |
| • NIVCD | 33 (9.7%) |
| • LAH | 25 (7.4%) |
| • Narrow QRS | 110 (32.4%) |
| • paced / escape / asystole | 52 (15.3%) |

RBBB – right bundle branch block; LAH - left anterior hemiblock, LPH - left posterior hemiblock; LBBB – left bundle branch block; NIVCD – non-specific intraventricular conduction delay.

A total of 1278 lead rotation events were analyzed with a median (IQR) of 3 (4) events per patient; in 25.9% of patients only one lead rotation event was necessary. Fixation beats were observed in 327 / 339 of final lead positions and in 9 / 939 intermediate lead positions; p < 0.001. The sensitivity, specificity, positive and negative predictive value of the LBB fixation beats as a marker for reaching the left bundle capture area was, 96.4%, 99.0%, 97.3% and 99.0%, respectively.

During the prospective phase of the study one lead rotation event was enough for 15 out of 21 patients (71.4%). The LBB fixation beats were observed in all 21 patients and only at LBB capture depth.

Pacing parameters and fluoroscopy time during the prospective phase did not differ when compared with the retrospective phase, while the number of lead rotation events lower - (Table 2).

**Table 2.** Pacing and procedure related characteristics

|  | Retrospective<br>n = 318 | Prospective<br>n = 21 | p |
| --- | --- | --- | --- |
| LBB capture confirmed with: |  |  |  |
| • Programmed stimulation | 241 (75.8%) | 17 (80.9%) | 0.59 |
| • Differential pacing output | 75 (23.9%) | 6 (28.6%) | 0.60 |
| • LBB potential | 111 (34.9%) | 11 (52.4%) | 0.11 |
| • Paced RWPT in V6 <85 ms | 176 (55.3%) | 18 (85.7%) | 0.006 |
| • Paced QRS < 120 ms* | 188 (59.1%) | 14 (66.7%) | 0.49 |
| • Just qR/Qr morphology in V1 | 11 (3.5%) | 0 (0%) | 1.0 |
| Paced QRS duration*[ms] | 151.7 ± 20.5 | 145.7±21.7 | 0.19 |
| Ventricular sensing [mV] | 8.5 ± 3.0 | 9.3 ± 3.9 | 0.36 |
| Acute LBB capture threshold [V] | 0.8 ± 0.5 | 0.9 ± 0.4 | 0.65 |
| Fluoroscopy time [min] | 11.4 ± 11.0 | 7.6 ± 5.3 | 0.12 |
| Lead rotation episodes [median (IQR)] | 3 (4) | 1 (1) | < 0.001 |
| Acute perforation to LV cavity [n] | 6 | 0 | 1.0 |
| Type of implanted device: |  |  | 0.51 |
| • DDD/VVI pacemaker | 249 (78.3%) | 17 (80.9%) | 0.39 |
| • CRT device | 69 (21.7%) | 4 (91.0%) |  |

LBB – left bundle branch; RWPT – R wave peak time; LV – left ventricle; CRT – cardiac resynchronization therapy; IQR – interquartile range
\* Measured from real QRS onset rather than from pacing spike

### Relationship between fixation beats QRS and paced QRS morphology

Pacing from the 3830 lead after observation of LBB area fixation beats, resulted in very similar/identical paced QRS as the LBB area fixation beats that preceded the pace-mapping in 325 out of 327 cases (99.4%). In several cases, a ‘shower’ of fixation beats with a progressive change of QRS was observed - that closely resembled progressive changes of QRS morphology that can be appreciated when constant pace-mapping is performed during lead deployment. In 136 / 339 (40.1%) patients intermediate beats – ‘deep septal fixation beats’ were observed, that often closely resembled paced morphology obtained at that particular intermediate lead depth in the interventricular septum (Figure 4 and supplemental Figure 2).

**Figure 4.**
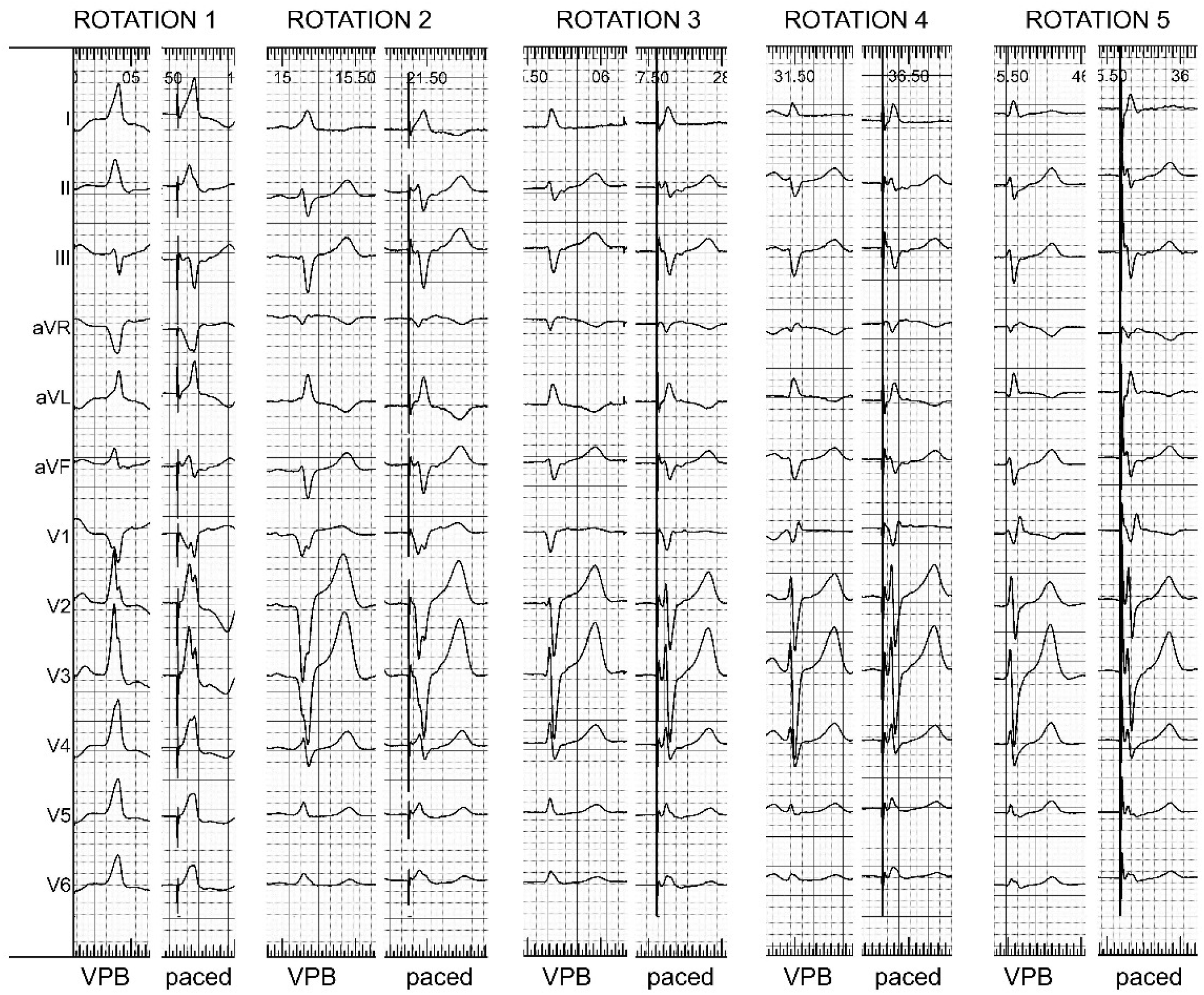
The fixation beats (VPB) obtained during 5 separate, consecutive lead rotation episodes in the same patient had very similar morphology as the paced QRS complexes obtained at the same site/depth in the interventricular septum. VPB – premature ventricular beat

### Perforations

In 6 patients included in the retrospective phase evident acute perforation to the left ventricle occurred. In the last one of these cases it was possible to appreciate the relation with LBB area fixation beats: lead rotations were continued despite occurrence of this marker, soon a shower of ectopics resulting from free movement of the pacing lead inside the left ventricular cavity was observed indicating that perforation occurred. These ‘perforation beats’ although also of right bundle branch type morphology where distinctly different than the deep septal ectopics resulting from the irritation of the LBB area (Figure 5). During the prospective phase no obvious perforations were noted and one microperforation was suspected when the rule was broken and a ‘bonus’ 1-2 lead rotation were added (looking for a bigger LBB potential) after the LBB area fixation beats were already observed. Microperforation was suspected on the basis of rise of capture threshold from the tip electrode and low capture threshold with exceptionally narrow QRS when pacing from the ring electrode. The lead was repositioned to a different location, this time stopping rotations immediately after the first LBB area fixation beat.

**Figure 5.**
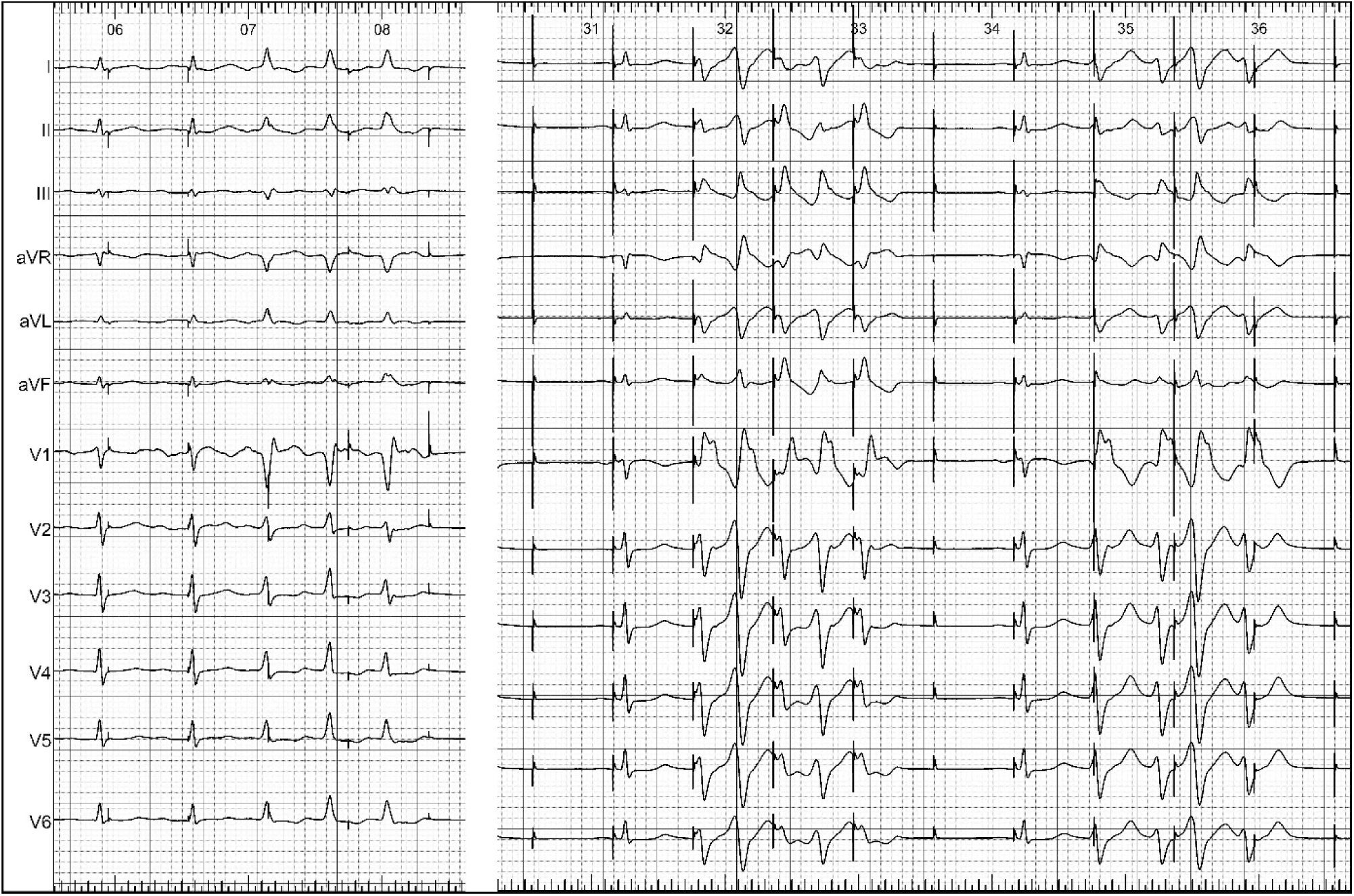
In the left panel LBB area fixation beats appeared during lead rotation for deep septal lead deployment. Further lead rotation resulted in perforation and salvos of left ventricular ectopic beats - illustrated in the right panel. Note the differences in QRS morphology between the LBB area fixation beats and the perforation beats; the last one show dominant R in V1 with notch on the descending limb of the R wave (Marriott’s sign) and wide QRS of 190 ms, while the former have typical Qr configuration in V1 and QRS duration does not exceed 125 ms.

## Discussion

The primary finding of our study is that the LBB fixation beats are occurring almost always when the deep septal lead approaches the left bundle branch capture area and almost never before. Consequently, they can serve as a potent, very specific marker during left bundle branch pacing lead implantation, a marker that indicates that LBB capture area was already reached and lead rotations should be immediately stopped to prevent perforation.

### Clinical translation

Deep septal deployment of the pacing lead is one of the crucial aspects of the LBB pacing technique. Monitoring the lead behavior and the lead depth in the septum is necessary to achieve LBB capture and at the same time to avoid perforation into the left ventricular cavity. Recent documents concerning practical aspects of LBB pacing proposed six measures to monitor lead depth: ‘fulcrum sign’, sheath angiography, impedance monitoring, changes in the notch in V1 lead, pacing from the ring electrode and monitoring of the endocardial signal.

However, of these, the fulcrum sign, sheath angiography, pacing from the ring electrode and impedance are only rough estimates of the lead depth and cannot provide real-time information that LBB capture area was reached. Pacing from the ring electrode is relatively more informative that the other methods as paced QRS morphology is a good indicator of pacing site within the septum. Moreover, these methods cannot prevent perforation as it can occur at an unpredictable lead depth due to the variable thickness of the interventricular septum and due to the variable course of the lead (oblique vs. perpendicular).

Monitoring changes in the notch in V1 lead is very useful but limited by the fact that a substantial percentage of patients do not have a notch in lead V1 at the initial site. Monitoring paced QRS morphology in all 12-leads leads seems superior, since there is a progressive change of the paced QRS morphology in all 12-leads, as the pacing lead gets deeper and deeper in the septum. Moreover, LBB-capture paced QRS, apart from qR/rsR’ morphology in V1, has several other typical features that enable to determine if LBB capture area was reached – for example global QRS duration, lack of QRS notch/slur or time to R wave peak in lead V6. Early in the development of the LBB pacing technique we have proposed the method of constant pacemapping while screwing the pacing lead in. ^9, 10^ That seemed be the perfect method as it enabled to see continuous change in paced QRS morphology and to terminate the lead fixation immediately when a satisfactory paced QRS was obtained. However, the lack of commercially available revolving connectors for the distal pin of the 3830 lead and the fragile, prone to break, connection between the distal pin and the inner conducting wire of that lead limited practical application of this technique. Therefore, the current mainstay technique of paced QRS morphology monitoring is the interrupted pacing method. The distal lead pin (cathode) has to be disconnected from the external pacemaker for the period of lead fixation. Only after an operator-dependent number of lead rotations, the deep septal pacing is repeated to check if the lead has already reached the LBB area. This approach has some drawbacks: procedure prolongation, loss of torque in the lead as the rotations are stopped, risk of sheath displacement or loss of proper sheath support, and the risk that the optimal lead depth might be missed and the lead would be already too deep when pace-mapping is performed.

The currently analyzed method of fixation beats monitoring for determination of lead depth and reaching LBB area is closely related to the monitoring of paced QRS. We believe the ectopic beats induced mechanically, as the lead crosses the septum, do not differ in terms of QRS morphology and diagnostic value from paced QRS. Not uncommonly it was possible to observe the whole spectrum of QRS morphologies of mechanically induced ectopic beats: from right ventricular septal beats, through deep septal beats, to left LBB area beats (Figure 1). However, the LBB area seems more vulnerable to mechanical irritation than the deep septum as we have observed deep septal fixation beats only in 41% of patients while LBB fixation beats were present in 96% in retrospective phase – when we were not monitoring them, and in 100% of cases in the prospective phase. This method has several advantages, firstly, it does not require connection of the pacing lead to the external pacemaker. Secondly, in vast majority of patients it is possible to reach LBB capture area with single lead rotation episode, using the same momentum. This not only speeds the procedure but possibly prevents dislodgements which in our experience are caused mainly by the drill effect (rotation of the lead in the same position without forward movement of the helix). Thirdly, this technique has the capacity to prevent perforations, especially microperforations. We could not accurately assess the frequency of microperforations during the retrospective phase. However, in our experience microperforations using the interrupted pacemapping method were occasionally observed by all operators.

### Limitations

The prospective phase of the study included relatively small number of patients. Albeit it provided very consistent results and fully compatible with the retrospective phase. The whole study was conducted in a single, university-based center with an experience of nearly one thousand conduction system implantations. It is possible that during rapid advancement of the lead in a non-LBB area, in the absence of fixation beats, LV septal perforation can occur. In patients with underlying RBBB, monitoring for LBB-fixation beats can be challenging. Vigilant monitoring for the occurrence of LBB-fixation beats is of paramount importance to avoid perforation. Fixation beats inform about reaching LBB capture area but are not diagnostic in itself of LBB capture. LBB capture still needs to be determined on the basis of criteria and maneuvers - as when this area is reached using the interrupted pacing method.

## Conclusion

This study analyzed the largest cohort of LBB paced patients to-date. The results suggest that monitoring 12-lead ECG for the occurrence of fixation beats during deep septal lead deployment might be an important addition to the LBB implantation technique. It has the potential to facilitate the procedure and may increase the safety of deep septal lead implantation. Multicenter prospective study seems warranted to further delineate usefulness of this implantation method.

## Data Availability

Data available upon reasonable request

## Figures and Legends

**Supplemental Figure 1.**
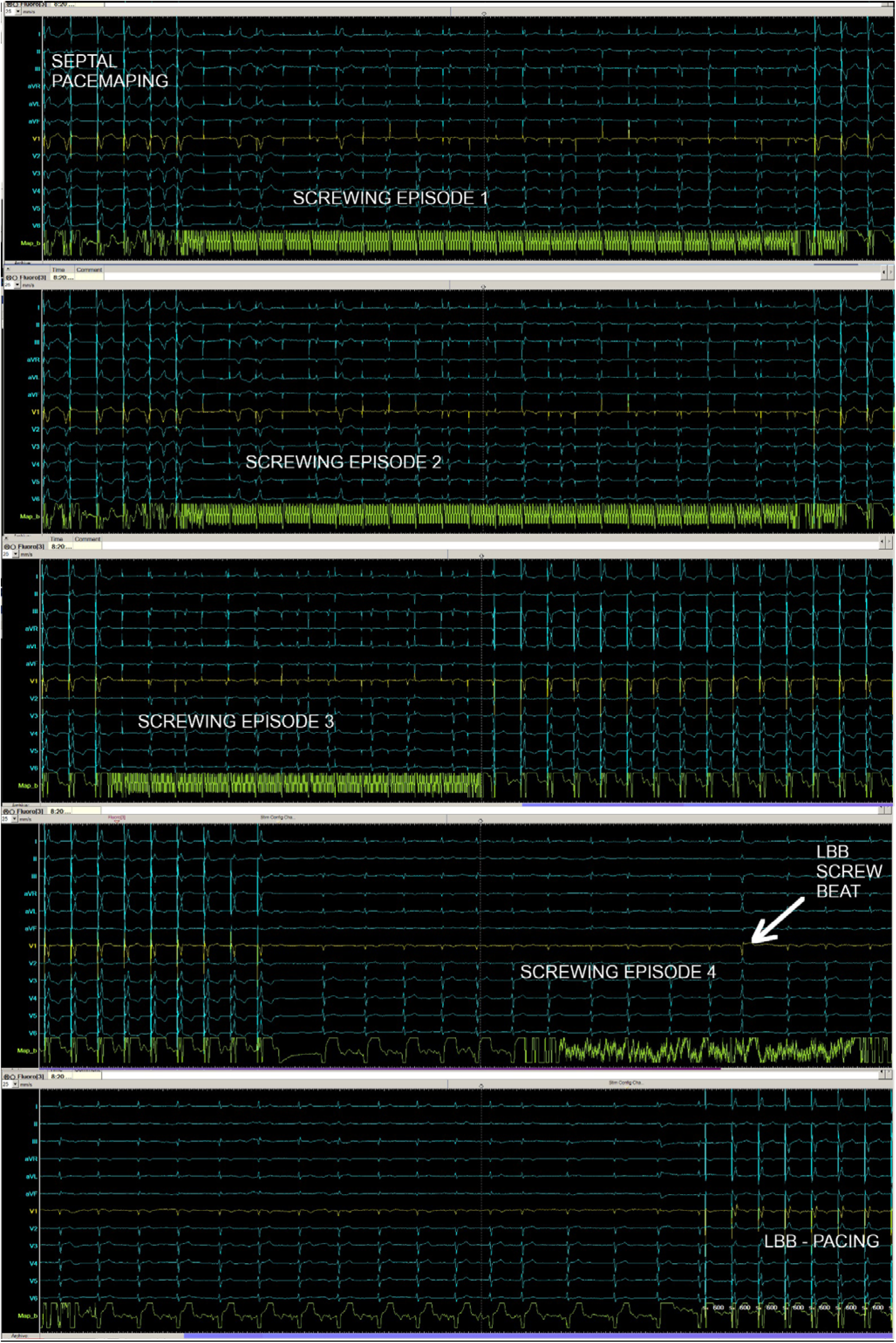
Continuous recording of 12-lead surface ECG and the endocardial electrogram from the 3830 pacing lead during deployment of the lead into the interventricular septum. Episodes of lead rotation are interspersed with pace-mapping to confirm lead position and capture of the left bundle branch. It took four lead rotation events to obtain a single LBB area fixation beat; after this, the pace-mapping revealed paced QRS morphology identical to fixation beat QRS morphology. LBB – left bundle branch.

**Supplemental Figure 2.**
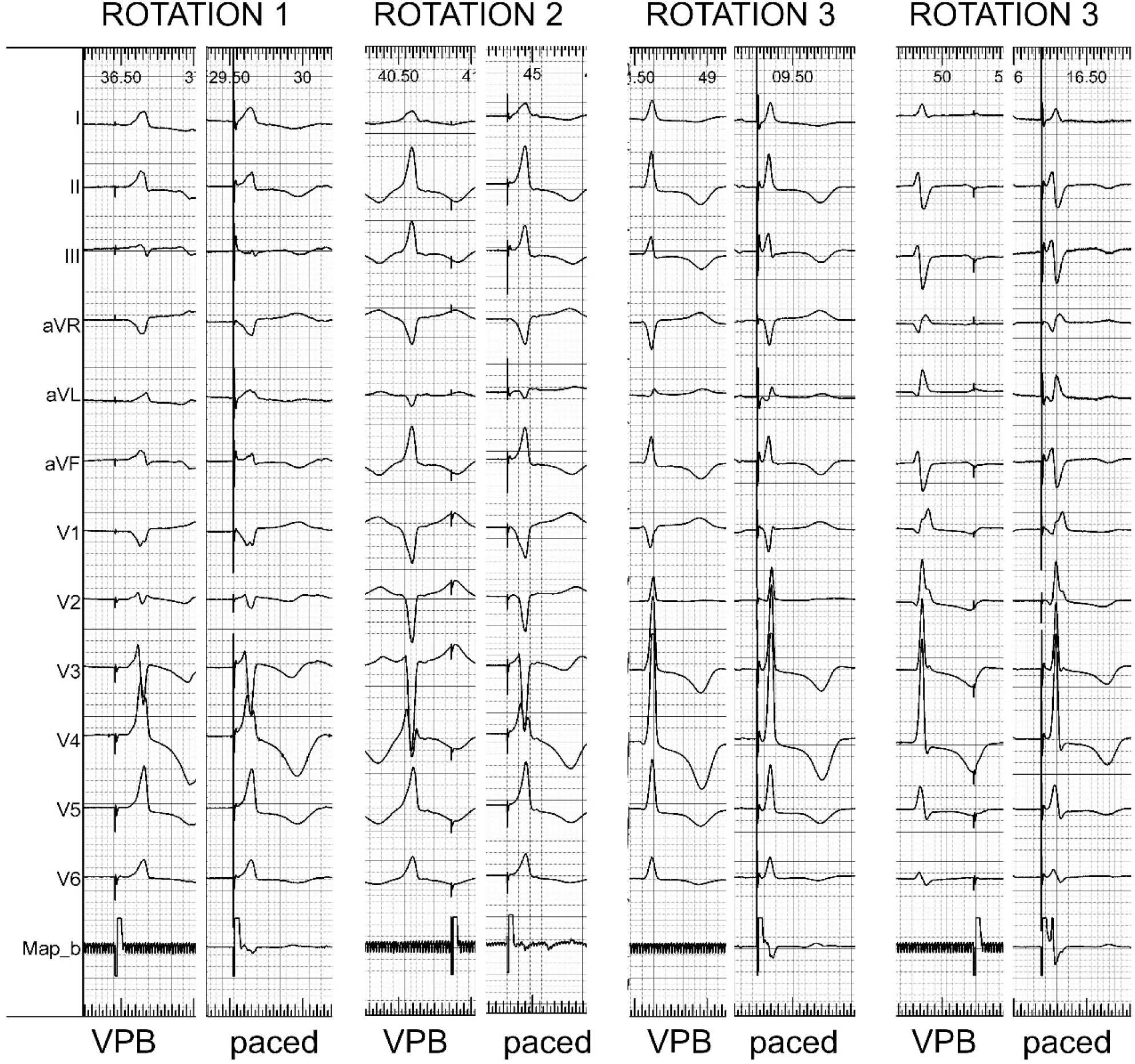
Fixation beats and corresponding paced QRS at the same site/depth in the interventricular septum. Note that, during the last lead rotation episode both LBB selective and LBB non-selective fixation betas were present, and both could be reproduced with pacing.

